# Predicting the cumulative number of cases for the COVID-19 epidemic in China from early data

**DOI:** 10.1101/2020.03.11.20034314

**Authors:** Z. Liu, P. Magal, O. Seydi, G. Webb

## Abstract

We model the COVID-19 coronavirus epidemic in China. We use early reported case data to predict the cumulative number of reported cases to a final size. The key features of our model are the timing of implementation of major public policies restricting social movement, the identification and isolation of unreported cases, and the impact of asymptomatic infectious cases.

## 1 Introduction

Many mathematical models of the COVID-19 coronavirus epidemic in China have been developed, and some of these are listed in our references [4, 7, 9, 10, 11, 12, 13, 14, 15]. We develop here a model describing this epidemic, focused on the effects of the Chinese government imposed public policies designed to contain this epidemic, and the number of reported and unreported cases that have occurred. Our model here is based on our model of this epidemic in [5], which was focused on the early phase of this epidemic (January 20 through January 29) in the city of Wuhan, the epicenter of the early outbreak. During this early phase, the cumulative number of daily reported cases grew exponentially. In [5], we identified a constant transmission rate corresponding to this exponential growth rate of the cumulative reported cases, during this early phase in Wuhan.

On January 23, 2020, the Chinese government imposed major public restrictions on the population of Wuhan. Soon after, the epidemic in Wuhan passed beyond the early exponential growth phase, to a phase with slowing growth. In this work, we assume that these major government measures caused the transmission rate to change from a constant rate to a time dependent exponentially decreasing rate. We identify this exponentially decreasing transmission rate based on reported case data after January 29. We then extend our model of the epidemic to the central region of China, where most cases occurred. Within just a few days after January 29, our model can be used to project the time-line of the model forward in time, with increasing accuracy, to a final size.

## 2 Model

The model consists of the following system of ordinary differential equations:

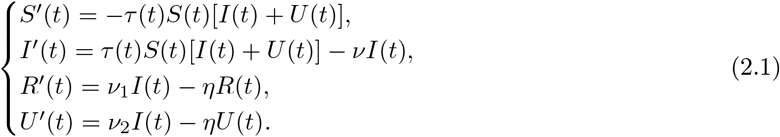

This system is supplemented by initial data

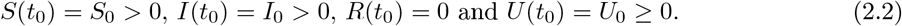

Here *t ≥ t*_0_ is time in days, *t*_0_ is the beginning date of the model of the epidemic, *S*(*t*) is the number of individuals susceptible to infection at time *t, I*(*t*) is the number of asymptomatic infectious individuals at time *t, R*(*t*) is the number of reported symptomatic infectious individuals at time *t*, and *U* (*t*) is the number of unreported symptomatic infectious individuals at time *t*.

Asymptomatic infectious individuals *I*(*t*) are infectious for an average period of 1*/ν* days. Reported symptomatic individuals *R*(*t*) are infectious for an average period of 1*/η* days, as are unreported symptomatic individuals *U* (*t*). We assume that reported symptomatic infectious individuals *R*(*t*) are reported and isolated immediately, and cause no further infections. The asymptomatic individuals *I*(*t*) can also be viewed as having a low-level symptomatic state. All infections are acquired from either *I*(*t*) or *U* (*t*) individuals.

The parameters of the model are listed in Table 1 and a schematic diagram of the model is given in Figure 1.

**Table 1:** Parameters of the model.

| Symbol | Interpretation | Method |
| --- | --- | --- |
| $t_0$ | Time at which the epidemic started | fitted |
| $S_0$ | Number of susceptible at time $t_0$ | fixed |
| $I_0$ | Number of asymptomatic infectious at time $t_0$ | fitted |
| $U_0$ | Number of unreported symptomatic infectious at time $t_0$ | fitted |
| $\tau(t)$ | Transmission rate at time $t$ | fitted |
| $1/\nu$ | Average time during which asymptomatic infectious are asymptomatic | fixed |
| $f$ | Fraction of asymptomatic infectious that become reported symptomatic infectious | fixed |
| $\nu_1 = f\nu$ | Rate at which asymptomatic infectious become reported symptomatic | fitted |
| $\nu_2 = (1-f)\nu$ | Rate at which asymptomatic infectious become unreported symptomatic | fitted |
| $1/\eta$ | Average time symptomatic infectious have symptoms | fixed |

**Figure 1:**
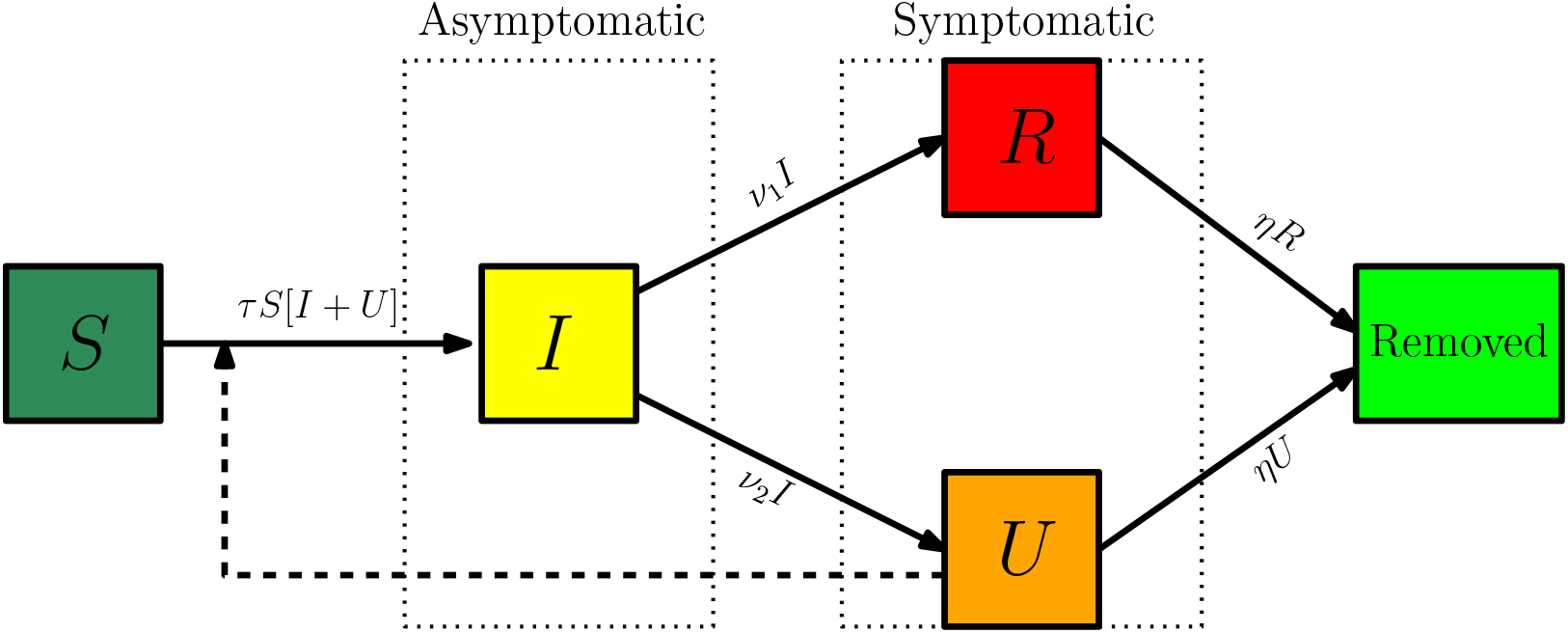
Compartments and flow chart of the model.

## 3 Data

We use cumulative reported data from the National Health Commission of the People’s Republic of China and the Chinese CDC for mainland China. Before February 11, the data was based on confirmed testing. From February 11 to February 15, the data included cases that were not tested for the virus, but were clinically diagnosed based on medical imaging showing signs of pneumonia. There were 17,409 such cases from February 10 to February 15. The data from February 10 to February 15 specified both types of reported cases. From February 16, the data did not separate the two types of reporting, but reported the sum of both types. We subtracted 17,409 cases from the cumulative reported cases after February 15 to obtain the cumulative reported cases based only on confirmed testing after February 15. The data is given in Table 2 with this adjustment..

**Table 2:** Cumulative daily reported case data from January 20, 2020 to February 15, 2020, reported for mainland China by the National Health Commission of the People’s Republic of China and the Chinese CDC. The data corresponds to cumulative reported cases confirmed by testing

| January |  |  |  |  |  |  |  |  |  |  |  |  |
| --- | --- | --- | --- | --- | --- | --- | --- | --- | --- | --- | --- | --- |
| 19 | 20 | 21 | 22 | 23 | 24 | 25 | 26 | 27 | 28 | 29 | 30 | 31 |
| 198 | 291 | 440 | 571 | 830 | 1287 | 1975 | 2744 | 4515 | 5974 | 7711 | 9692 | 11791 |
| February |  |  |  |  |  |  |  |  |  |  |  |  |
| 1 | 2 | 3 | 4 | 5 | 6 | 7 | 8 | 9 | 10 | 11 | 12 | 13 |
| 14380 | 17205 | 20438 | 24324 | 28018 | 31161 | 34546 | 37198 | 40171 | 42638 | 44653 | 46472 | 48467 |
| 14 | 15 | 16 | 17 | 18 | 19 | 20 | 21 | 22 | 23 | 24 | 25 | 26 |
| 49970 | 51091 | 53139 | 55027 | 56776 | 57593 | 58482 | 58879 | 59527 | 59741 | 60249 | 60655 | 61088 |
| 27 | 28 | 29 |  |  |  |  |  |  |  |  |  |  |
| 61415 | 61842 | 62415 |  |  |  |  |  |  |  |  |  |  |
| March |  |  |  |  |  |  |  |  |  |  |  |  |
| 1 | 2 | 3 | 4 | 5 | 6 |  |  |  |  |  |  |  |
| 62617 | 62742 | 62861 | 63000 | 63143 | 63242 |  |  |  |  |  |  |  |

We plot the data for daily reported cases and the cumulative reported cases in Figure 2.

**Figure 2:**
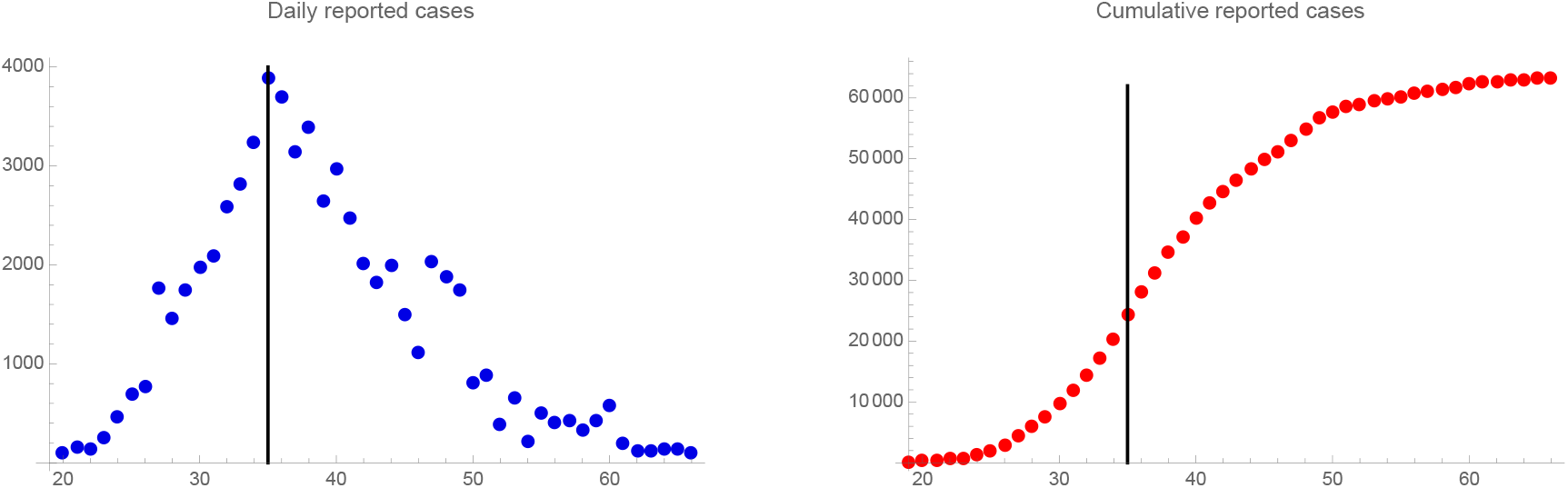
Daily reported cases data (left) and cumulative reported cases data (right). The epidemic turning point of the reported case data is approximately February 4, 2020 (day 35, day 1 = January 1, 2020).

## 4 Model parameters

We assume *f* = 0.8, which means that 20% of symptomatic infectious cases go unreported. We assume *η* = 1*/*7, which means that the average period of infectiousness of both unreported symptomatic infectious individuals and reported symptomatic infectious individuals is 7 days. We assume *ν* = 1*/*7, which means that the average period of infectiousness of asymptomatic infectious individuals is 7 days. These values can be modified as further epidemiological information becomes known.

A illustrated in our previous work [5], we assume that in the early phase of the epidemic, the cumulative number of reported cases grow approximately exponentially, according to the formula:

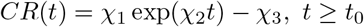

with values *χ*_1_ = 0.15, *χ*_2_ = 0.38, *χ*_3_ = 1.0. These values of *χ*_1_, *χ*_2_, and *χ*_3_ are fitted to reported case data from January 19 to January 28. We assumed the initial value *S*_0_ = 11, 000, 000, the population of the city Wuhan, which was the epicenter of the epidemic outbreak where almost all cases in China occurred in this time period. The other initial conditions are

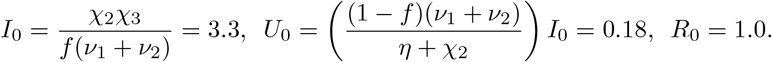

The value of the transmission rate *τ* (*t*), during the early phase of the epidemic, when the cumulative number of reported cases was approximately exponential, is the constant value

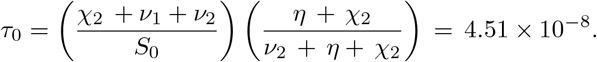

The initial time is

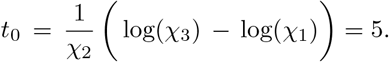

The value of the basic reproductive number is

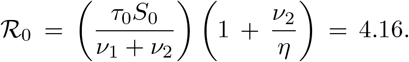

These parameter formulas were derived in [5].

After January 23, strong government measures in all of China, such as isolation, quarantine, and public closings, strongly impacted the transmission of new cases. The actual effects of these measures were complex, and we use an exponential decrease for the transmission rate *τ* (*t*) to incorporate these effects after the early exponentially increasing phase. The formula for *τ* (*t*) during the exponential decreasing phase was derived by a fitting procedure. The formula for *τ* (*t*) is

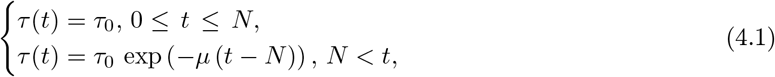

The date *N* and the value of *µ* are chosen so that the cumulative reported cases in the numerical simulation of the epidemic aligns with the cumulative reported case data during a period of time after January 19. We choose *N* = 25 (January 25) for our simulations. We illustrate *τ* (*t*) in Figure 3, with *µ* = 0.16. In this way we are able to project forward the time-path of the epidemic after the government imposed public restrictions, as it unfolds.

**Figure 3:**
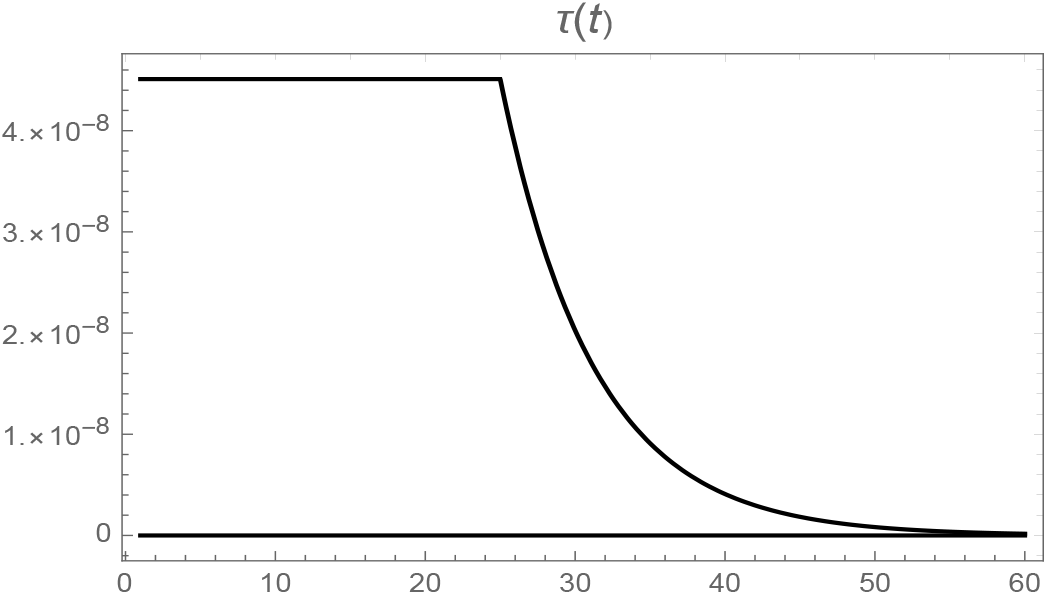
Graph of τ (t) with N = 25 (January 25) and µ = 0.16. The transmission rate is effectively 0.0 after day 53 (February 22).

## 5 Model simulation

We assume that exponentially increasing phase of the epidemic (as incorporated in *τ*_0_) is intrinsic to the population of any subregion of China, after it is has been established in the epidemic epicenter Wuhan. We also assume that the susceptible population *S*(*t*) in not significantly reduced over the course of the epidemic. We set *τ*_0_ = 4.51*×* 10^*−*8^, *t*_0_ = 5.0, *I*(*t*_0_) = 3.3, *U* (*t*_0_) = 0.18, and *R*(*t*_0_) = 1.0, as in Section 4. We set *S*(*t*_0_) in (2.2) to 1, 400, 050, 000 (the population of mainland China). We set *τ* (*t*) in (2.1) to

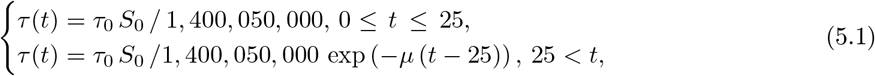

where *S*_0_ = 11, 000, 000 (the population of Wuhan). We thus assume that the government imposed restriction measures became effective in reducing transmission on January 25.

In Figure 4, we plot the graphs of *CR*(*t*), *CU* (*t*), *R*(*t*), and *U* (*t*) from the numerical simulation for simulations based on six time intervals for known values of the cumulative reported case data. For each of these time intervals, a value of *µ* is chosen so that the simulation for that time interval aligns with the cumulative reported case data in that interval. In this way, we are able to predict the future values of the epidemic from early cumulative reported case data.

**Figure 4:**
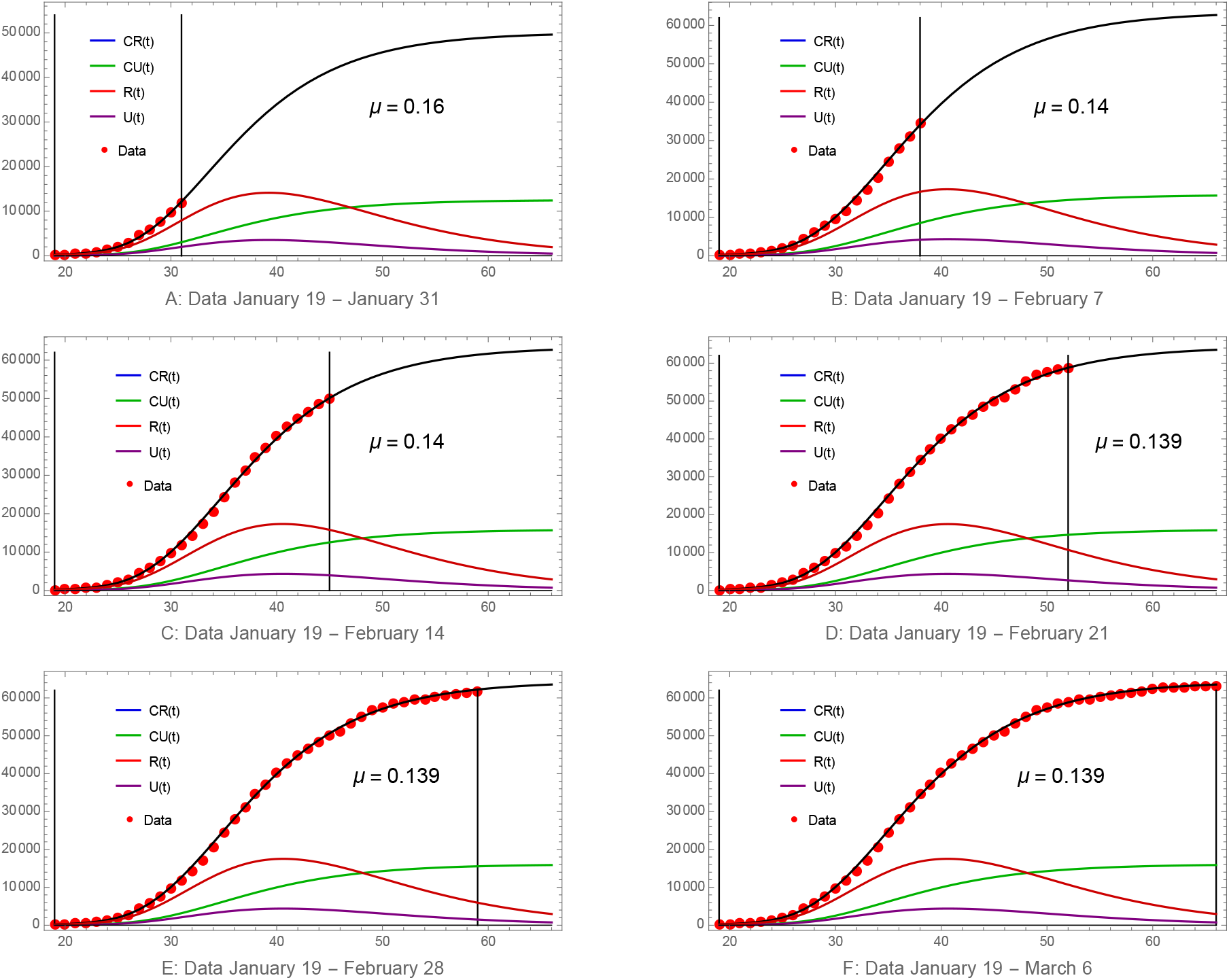
Graphs of CR(t), CU (t), U (t), R(t). The red dots are the reported case data. The value of µ (indicated in each of the sub-figures) is estimated by fitting the model output for each time interval to the cumulative reported case data for that time interval. The final size of cumulative cases is approximately (A) 49, 600 reported, 12, 400 unreported; (B) and (C) 62,700 reported, 15,700 unreported cases; and (D), (E), (F): 63, 500 reported, 15, 900 unreported.

In Figure 5 we plot the graphs of the reported cases *R*(*t*), the unreported cases *U* (*t*), and the infectious pre-symptomatic cases *I*(*t*). The blue dots are obtained from the reported cases data (Table 2) for each day beginning on January 26, by subtracting from each day, the value of the reported cases one week earlier.

**Figure 5:**
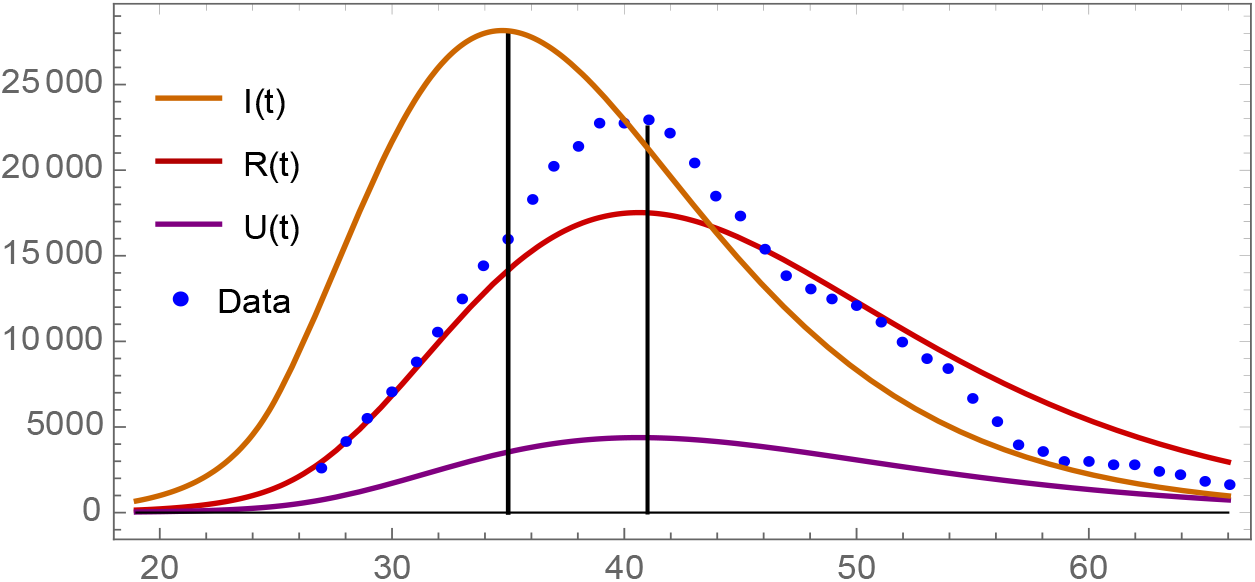
Graphs of R(t), U (t), I(t). The blue dots are the day by day weekly reported data. The turning point of the asymptomatic infectious cases I(t) is approximately day 35 = February 4. The turning point of the reported cases R(t) and the unreported cases U (t) is approximately day 41 = February 10. The turning point of the day by day weekly reported data is approximately day 41.

Our model transmission rate *τ* (*t*) can be modified to illustrate the effects of an earlier or later implementation of the major public policy interventions that occurred in this epidemic. The implementation one week earlier (25 is replaced by 18 in (4.1)) is graphed in Figure 6A. All other parameters and the initial conditions remain the same. The total reported cases is approximately 4, 500 and the total unreported cases is approximately 1, 100. The implementation one week later (25 is replaced by 32 in (4.1)) is graphed in Figure 6B. The total reported cases is approximately 820, 000 and the total unreported cases is approximately 200, 000. The timing of the institution of major social restrictions is critically important in mitigating the epidemic.

**Figure 6:**
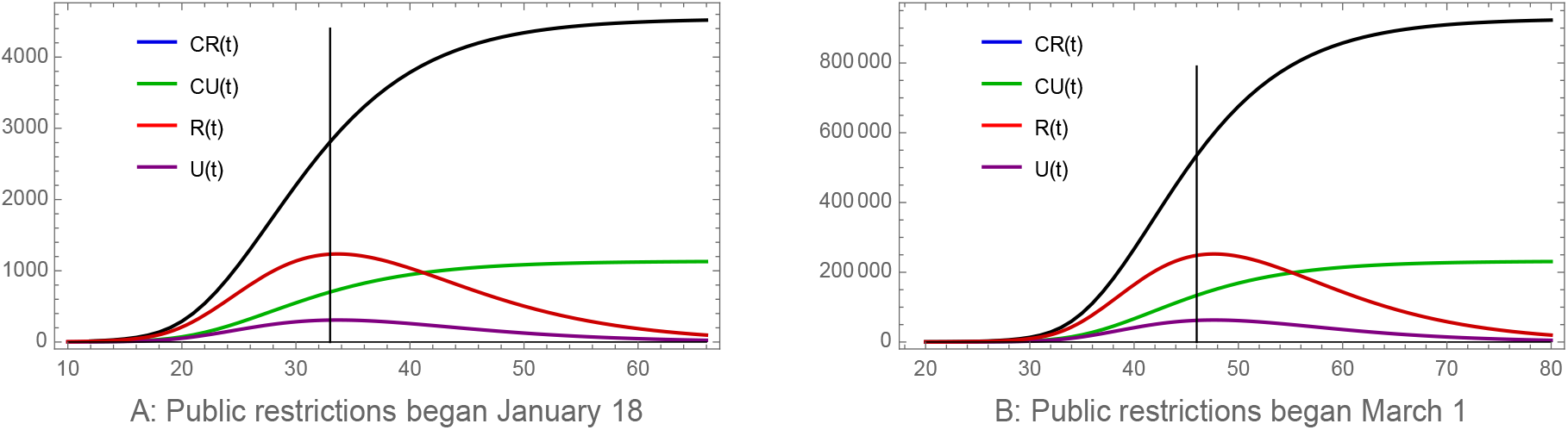
Graphs of CR(t), CU (t), U (t), R(t).A: The major public policy interventions were implemented one week earlier (January 18). B: The major public policy interventions were implemented one week later (March 1). The one week earlier turning point day is 33 = February 2. The one week later turning point is day 46 = February 15.

The number of unreported cases is of major importance in understanding the evolution of an epidemic, and involves great difficulty in their estimation. The data from January 19 to February 15 for reported cases in Table 2, was only for confirmed tested cases. Between February 11 and February 15, additional clinically diagnosed case data, based on medical imaging showing signs of pneumonia, was also reported by the Chinese CDC. Since February 16, only tested case data has been reported by the Chinese CDC, because new NHC guidelines removed the clinically diagnosed category. Thus, after February 15, there is a gap in the reported case data that we used up to February 15. The uncertainty of the number of unreported cases for this epidemic includes this gap, but goes even further to include additional unreported cases.

We assumed previously that the fraction *f* of reported cases was *f* = 0.8 and the fraction of unreported cases was 1 − *f* = 0.2. Our model formulation can be applied with varying values for the fraction *f*. In Figure 7, we provide illustrations with the fraction *f* = 0.4 (Figure 7A) and *f* = 0.6 (Figure 7B). For *f* = 0.4, the formula for the time dependent transmission rate *τ* (*t*) in (4.1) involves new values for *τ*_0_ = 4.08 *×* 10^*−*8^ and *µ* = 0.148. The initial conditions *I*_0_ = 6.65 and *U*_0_ = 1.09 also have new values. The other parameters and initial conditions remain the same. For *f* = 0.4, the total reported cases is approximately 63, 100 and the total unreported cases is approximately 94, 700. For *f* = 0.6, the formula for the time dependent transmission rate *τ* (*t*) in (4.1) involves new values for *τ*_0_ = 4.28 *×* 10^*−*8^ and *µ* = 0.144. The initial conditions *I*_0_ = 4.43 and *U*_0_ = 0.485 also have new values. The other parameters and initial conditions remain the same. For *f* = 0.6, the total reported cases is approximately 63, 300 and the total unreported cases is approximately 42, 200. From these simulations, we see that estimation of the number of unreported cases has major importance in understanding the severity of this epidemic.

**Figure 7:**
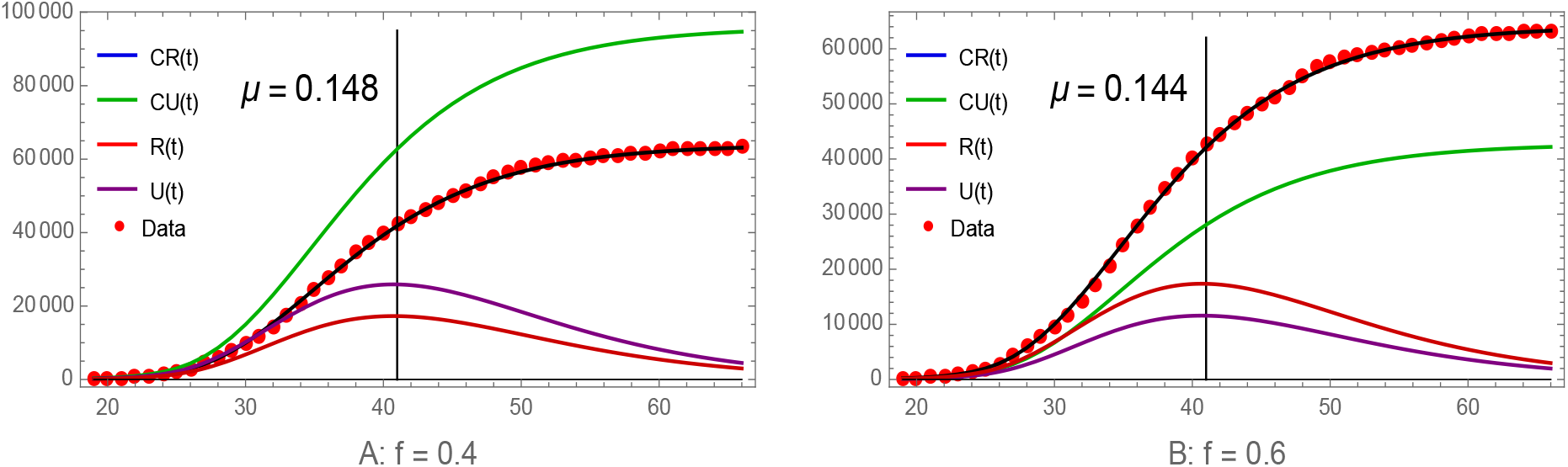
Graphs of CR(t), CU (t), U (t), R(t). The red dots are the cumulative reported case data from Table 2. A: f = 0.4. The basic reproductive number is ℛ_0_ = 5.03. The final size of the epidemic is approximately 157, 800 total cases. The turning point of the epidemic is approximately day 41 = February 10. B: f = 0.6, The basic reproductive number is ℛ_0_ = 4.62. The final size of the epidemic is approximately 105, 500 total cases. The turning point is approximately day 41 = February 10.

The number of days an asymptomatic infected individual is infectious is uncertain. We simulate in Figure 8 the model with *ν* = 1*/*3 (asymptomatic infected individuals are infectious on average 3 days before becoming symptomatic), and *ν* = 1*/*5 (asymptomatic infected individuals are infectious on average 5 days before becoming symptomatic). For *ν* = 1*/*3, *τ*_0_ = 5.75 *×* 10^*−*8^, *I*_0_ = 1.48, *U*_0_ = 0.18, *R*_0_ = 2.78, *µ* = 0.0765, *N* = 24. For *ν* = 1*/*5, *τ*_0_ = 4.90 *×* 10^*−*8^, *I*_0_ = 2.38, *U*_0_ = 0.18, *R*_0_ = 3.45, *µ* = 0.098, *N* = 24. Because the asymptomatic infectious periods are shorter, The date *N* of the effective reduction of the transmission rate due to restriction measures, is one day earlier.

**Figure 8:**
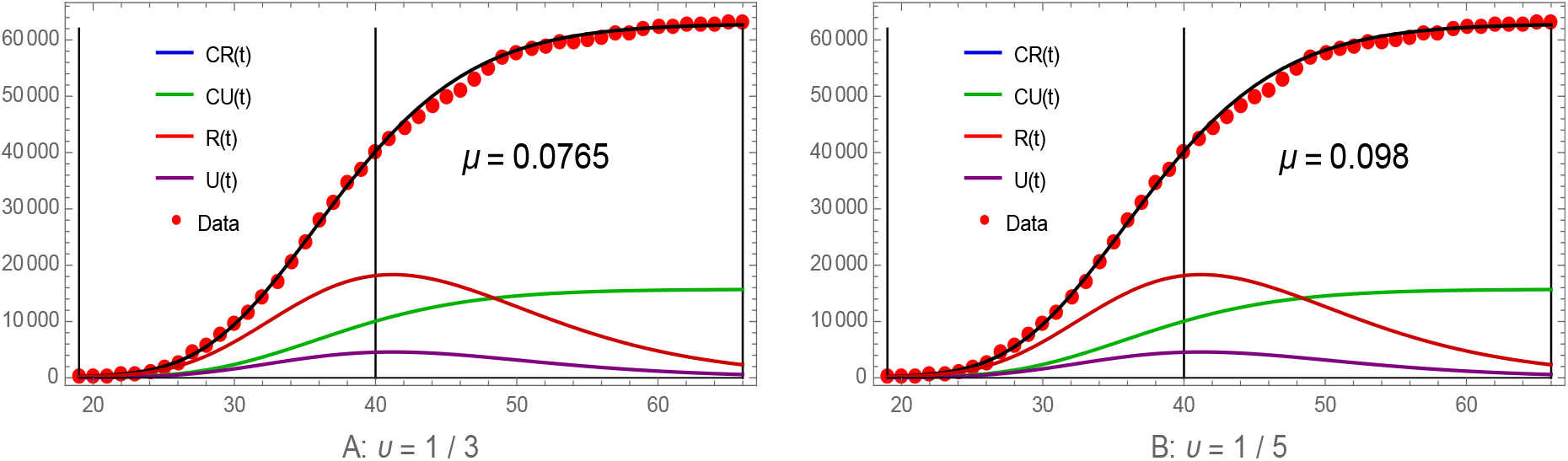
Graphs of CR(t), CU (t), U (t), R(t). The red dots are the cumulative reported case data from Table 2. A: ν = 1/3. B: ν = 1/5. The turning point is approximately day 40 = February 9 for both cases. The final sizes of CR(t) and CU (t) are approximately the same as for ν = 1/7 in both cases.

In Figure 9, we illustrate the importance of the level of government imposed public restrictions by altering the value of *µ* in formula (4.1). All other parameters and initial conditions are the same as in Figure 4. In Figure 9A we set *µ* = 0.0, corresponding to no restrictions. The final size of cumulative reported cases after 100 days is approximately 1,080,000,000 cases, approximately 270,000,000 unreported cases, and approximately 1,350,000,000 total cases. The turning point is approximately day 65 = March 6. In Figure 9B we set *µ* = 0.17, corresponding to a higher level of restrictions than in Figure 4. The final size of cumulative reported cases after 70 days is approximately 45,300 cases, approximately 11,300 unreported cases, and approximately 56,600 total cases. The turning point is approximately day 38 = February 7. The level and timing of government restrictions on social distancing is very important in controlling the epidemic.

**Figure 9:**
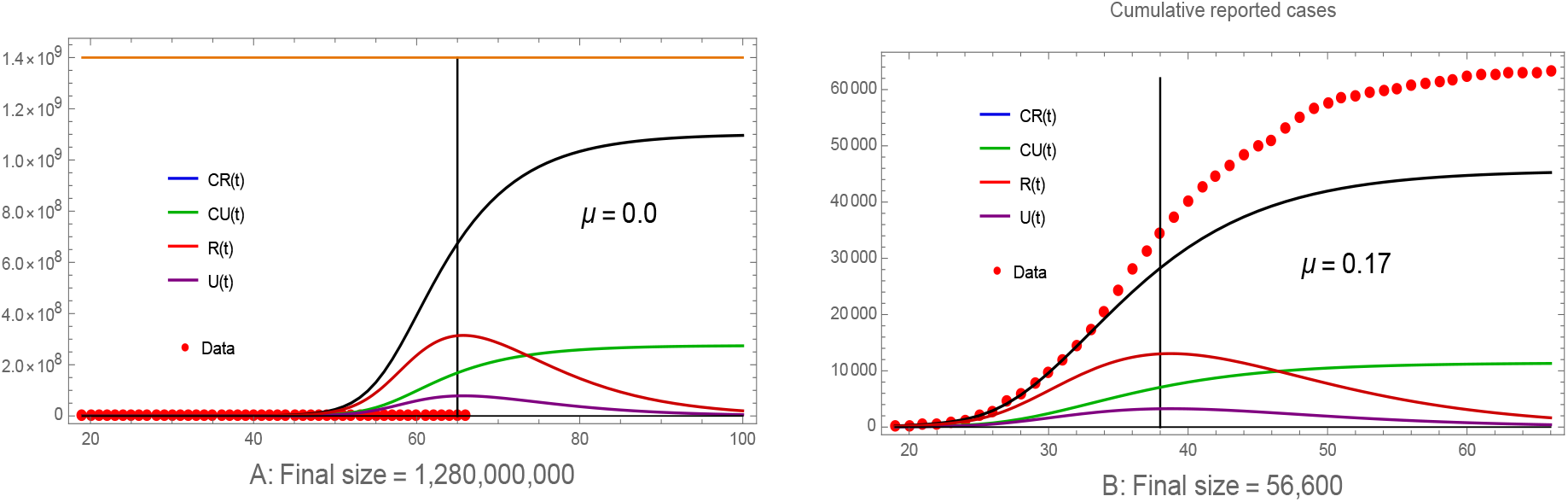
Graphs of CR(t), CU (t), U (t), and R(t). case data from Table 2. A: µ = 0.0 The orange horizontal line is the population of mainland China. B: µ = 0.17. A significant reduction of cases occurs compared to the cumulative reported case data (red dots).

## 6 Discussion

We have developed a model of the COVID-19 epidemic in China that incorporates key features of this epidemic: (1) the importance of the timing and magnitude of the implementation of major government public restrictions designed to mitigate the severity of the epidemic; (2) the importance of both reported and unreported cases in interpreting the number of reported cases; and (3) the importance of asymptomatic infectious cases in the disease transmission. In our model formulation, we divide infectious individuals into asymptomatic and symptomatic infectious individuals. The symptomatic infectious phase is also divided into reported and unreported cases. Our model formulation is based on our work [5], in which we developed a method to estimate epidemic parameters at an early stage of an epidemic, when the number of cumulative cases grows exponentially. The general method in [5], was applied to the COVID-19 epidemic in Wuhan, China, to identify the constant transmission rate corresponding to the early exponential growth phase.

In this work, we use the constant transmission rate in the early exponential growth phase of the COVID-19 epidemic identified in [5]. We model the effects of the major government imposed public restrictions in China, beginning on January 23, as a time-dependent exponentially decaying transmission rate after January 24. With this time dependent exponentially decreasing transmission rate, we are able to fit with increasing accuracy, our model simulations to the Chinese CDC reported case data for all of China, forward in time from February 15, 2020.

Our model demonstrates the effects of implementing major government public policy measures. By varying the date of the implementation of these measures in our model, we show that had implementation occurred one week earlier, then a significant reduction in the total number of cases would have resulted. We show that if these measures had occurred one week later, then a significant increase in the total number of cases would have occurred. We also show that if these measures had been less restrictive on public movement, then a significant increase in the total size of the epidemic would have occurred. It is evident, that control of a COVID-19 epidemic is very dependent on an early implementation and a high level of restrictions on public functions.

We varied the fraction 1*− f* of unreported cases involved in the transmission dynamics. We showed that if this fraction is higher, then a significant increase in the number of total cases results. If it is lower, then a significant reduction occurs. It is evident, that control of a COVID-19 epidemic is very dependent on identifying and isolating symptomatic unreported infectious cases. We also decreased the parameter *ν* (the reciprocal of the average period of asymptomatic infectiousness), and showed that the total number of cases in smaller. It is also possible to decrease *η* (the reciprocal of the average period of unreported symptomatic infectiousness), to obtain a similar result. It is evident that understanding of these periods of infectiousness is important in understanding the total number of epidemic cases.

Our model was specified to the COVID-19 outbreak in China, but it is applicable to any outbreak location for a COVID-19 epidemic.

## Data Availability

The data used are public (WHO and CDC)

